# Act early, save lives: managing COVID-19 in Greece

**DOI:** 10.1101/2020.04.29.20084798

**Authors:** Gountas Ilias, Hillas Georgios, Souliotis Kyriakos

## Abstract

**Objectives:** To assess the impact of the implemented social distancing interventions (SD) in Greece.

**Study Design:** A dynamic, discrete time, stochastic individual-based model was developed to simulate COVID-19 transmission.

**Methods:** We fit the transmission model to the observed trends in deaths and ICU beds use.

**Results:** If Greece had not implemented the SD measures, the healthcare system would have been overwhelmed between 30 March and 4 of April. Additionally, the SD interventions averted 4360 deaths and prevent the healthcare system from overwhelmed.

**Conclusions:** The fast reflexes of the Greek government limit the burden of the Covid-19 outbreak.

## Main text

### Introduction

The COVID-19 pandemic represents a global public health emergency, with over 2.4 million reported cases^1^. The absence of a vaccine or a targeted antiviral treatment led countries’ public health responses to be directed mostly to non-pharmaceutical interventions, like social distancing (SD) measures, to reduce the impact of the outbreak. The goal is to slow the spread of the virus in order to keep the number infected at a manageable level and ensure that hospitals would not be overwhelmed.

Greece is a country with a vulnerable public healthcare system since the footprints of the substantial financial recession of the previous decade are still visible. For example, the number of Intensive Care Unit (ICU) beds in Greece is significantly lower than the European average (about 6 ICU beds per 100.000 individuals; 47.8% lower than the European average)^2,3^. On 18 March the Ministry of Health announced in the Greek parliament that only 120 ICUs beds were available for the COVID-19 patients^4^. Given the above complexities, the country was at risk of facing a significant COVID-19 epidemic, which would strain the healthcare system.

The first laboratory-confirmed case and the first death were reported on 26 February and 12 March 2020, respectively. It is important that the SD interventions launched prior to the first death. More specifically, on 9, 16, and 19 March, schools/universities closed, social distancing was encouraged and public events banned, respectively. Four days later significant restrictions on public movements and gatherings were imposed. Thanks to the quick implementation of SD interventions, the deaths and the of imports in ICUs beds were kept in manageable levels^5^.

Public health decision-makers need evidence-based information to evaluate and update strategic interventions to minimize the COVID-19 outbreak. Mathematical models can provide important insights by examining the effectiveness of the already applied interventions^6^. The aim of this study is to disentangle the impact of the SD interventions implemented in Greece and to examine what would be the expected deaths and needs of the healthcare system if interventions had not been implemented.

## Methods

### Study Design

In infectious disease epidemiology, mathematical models are a common way to explain the spread of diseases and to predict/assess the impact of potential intervention policies. Those models stratify individuals into compartments representing different states of the infection process (e.g. susceptible, infected, recovered individuals). In those models, individuals are moving between states according to transition rates. The impact of an intervention can be simulated by modifying these rates, e.g. the effect of the SD measures can be simulated by reducing the probability of an individual becoming infected.

### Description of the mathematical model

To model COVID-19 transmission, a discrete-time, stochastic, individual-based model which categorizes the population into susceptible, exposed, infectious, or removed (SEIR) individuals was developed in C++ (Figure S1).

In short, every day a susceptible individual might acquire the infection enter the exposed disease state before they become infectious. According to the clinical experience in Greece, about 80–90% of the individuals would have no or mild symptoms and would not need hospitalization. Those individuals after 5–7 days would recover from the disease^7^. The rest of the population would need to enter the healthcare system. From them, the majority would need hospitalization, while a smaller group would need also to move to an ICU bed. The average duration that a patient needs to stay in the hospital provided he wouldn’t need transfer to an ICU bed is 15 days. If a patient needs to move to intensive care, he/she will stay there for 14 days and additional 14 days in the hospital for recovery. The probability of death for patients during hospitalization or in ICU bed is 15% and 50%, respectively^8^. Further details about the description of the model are available in the appendix.

### Model parameterization and examined scenarios

The model was calibrated using COVID-19 epidemiological and clinical data from the Greek epidemic. More specifically, we varied the transmission rate, the proportion of patients who would need to be hospitalized, and the effect of SD interventions to optimize the fit on the observed trends in deaths and ICU beds. The time horizon was set by 27 April, since this day is expected to be the last day of the existing social distancing measures.

A ‘status quo’ scenario was used to generate predictions regarding the observed course of the outbreak. In addition, a scenario where all SD interventions were removed was implemented (“counterfactual scenario”), to estimate how the outbreak would have unfolded if nothing had been done. For each scenario, 1000 runs were performed, and results were summarized. In order to include the appropriate uncertainty (stochastic variability), the 2.5 and 97.5 percentiles of simulations were also shown.

## Results

### Epidemic parameters

After the reconstruction of the observed data on mortality and ICU beds in Greece, the best estimate for the basic reproduction number was R_0_ =2.6. Furthermore, the model estimates that 8.5% and 4.5% of the population would need hospitalization and be treated in ICU beds, respectively, if infected.

### Model projections under base case and counterfactual scenario

Figure 1a and 1b show that status quo scenario captures accurately the overall trends in deaths and the use of ICU beds prevalence between 27^th^ of February and 15^th^ of April.

The strain on the healthcare system would be significantly higher without the SD interventions. More specifically, if Greece had not implemented the SD interventions, the healthcare system would have been overwhelmed between 30 March and 4 of April (Figure 1).

Under the base case scenario, the number of covid-19-related deaths is anticipated to be 142 (95% credible intervals (CrI): 120–170) by 27 April. On the contrary, under the counterfactual scenario, that is without any SD intervention, the projected covid-19 related number of deaths would be 860 (95% CrI: 720, 1020) by 27 April (Figure 1b). Furthermore, considering the potential additional deaths that could occur due to the non-availability of an ICU bed, when the healthcare system reaches its limits, the disease-related mortality would be significantly higher (assumed that ICU beds availability would be the same as the pre-outbreak levels). Specifically, following the healthcare system overload (i.e. no additional availability of ICU beds), if we assumed that 90% of those who would be in need of an ICU bed would die, the additional deaths by 27 April would be 3500 (95% CrI: 2500, 4400). Summing the above, the expected deaths without the implementation of any interventions (neither SD measures nor increase ICU capacity) would be 4360 by 27 April.

Concerning the total number of cases under the status quo scenario, the model predicted that about 10.000 (95% CrI: 8.500–11.500) cases would exist in Greece by 27 April. In the absence of SD measures, the corresponding numbers would have been 230.000 (95% CrI: 208.000–247.000) (Figure 1c).

Regarding the course of the outbreak, the model estimated that the number of infected individuals peaked in Greece on 24 March (Figure S3). Finally, the R_0_ on 27 April is expected to be 0.52.

## Discussion

We reproduced the Covid-19 epidemic in Greece using a mathematical transmission model. To evaluate the impact of interventions, we fit a counterfactual model, without the SD measures, and compare this to the actual model. Our analysis highlights that SD interventions that took place early enough in the outbreak were highly successful, as they managed to keep the number of deaths and needed ICU beds at low levels, and within the capacity of the national healthcare system. It is estimated that the SD measures averted about 4360 deaths by 27 April. Additionally, our model highlights that any interventions to boost ICU capacity, without the simultaneous implementation of SD interventions, is inefficient healthcare policy, as the demand for those beds would be very high without applying the SD interventions.

It should be noted that our estimates regarding the effectiveness of the SD interventions are conservative, as in the counterfactual scenario we only removed the SD interventions while assuming that everything else would remain exactly the same as in the base case scenario. This assumption ignores additional potential complications, due to the healthcare system overload or collapse. When the healthcare system overwhelmed, several things would have been unfolded differently. For example, if the prevalence of COVI-19 inside the healthcare settings had increased, a larger number of doctors or nurses could be infected with the virus, and thus the capacity of the healthcare system would be reduced.

It is important that the outputs of our model have been also computed/observed elsewhere. Our estimates regarding the basic reproduction number and the total infected population compare well to the estimates from the Imperial College Covid19 Response Team^9^. Furthermore, It has been shown that patients who are most at risk to experience COVID-19 complications are elderly individuals^6^. In Greece, the proportion of population aged above 70 is 14.8%^10^, which is close to the estimates of our model that 13.0% of the population would need to enter the healthcare system if infected.

## Conclusions

The fast reflexes of the Greek government, which launched early enough the SD measures, managed to minimize the burden of the Covid-19 outbreak and prevent the overload of the healthcare system.

## Data Availability

All relevant data are within the paper

---

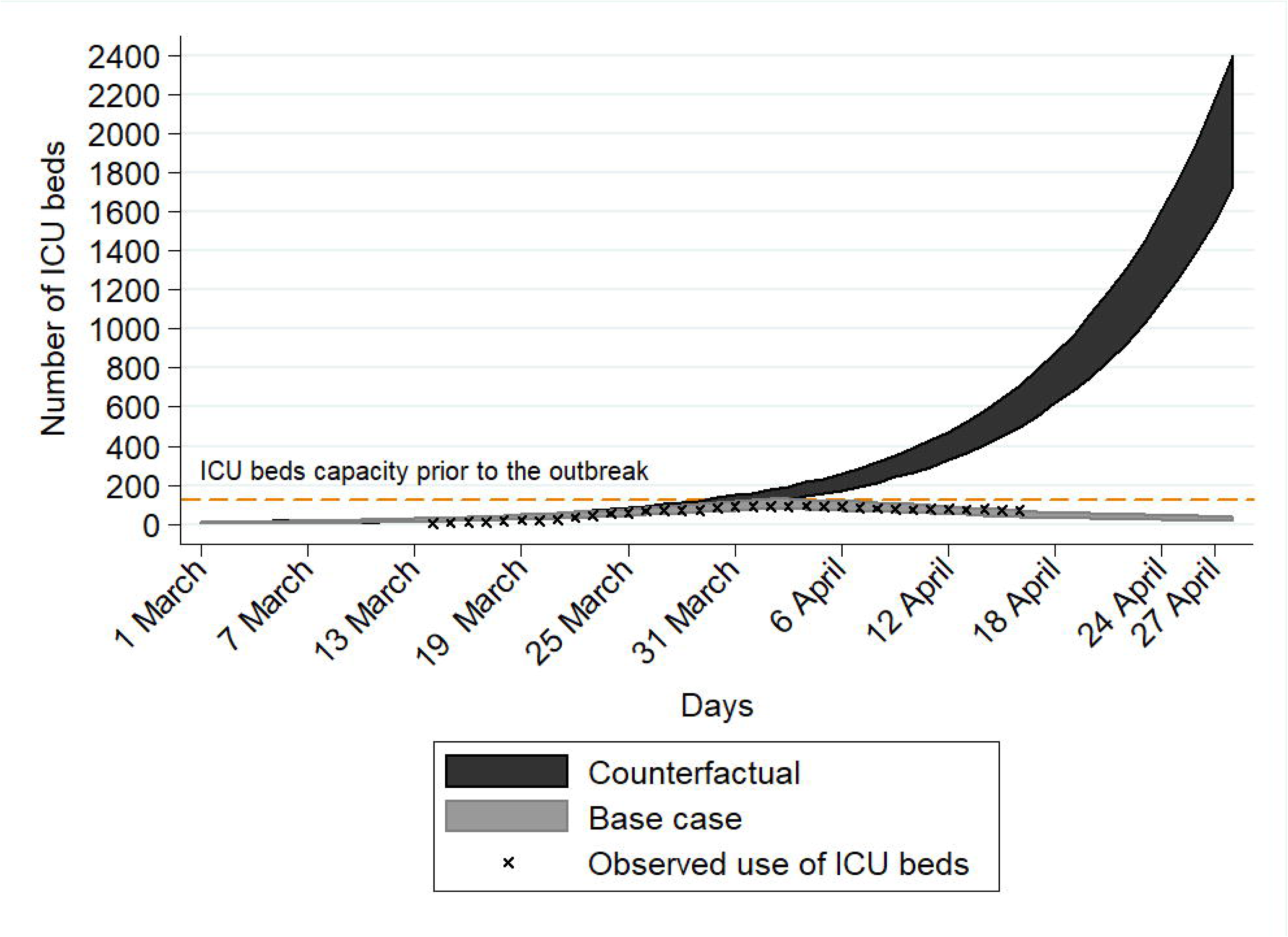
Figure: Projections of future COVID-19 cases and complications under status quo and counterfactual scenario. For comparison, x indicates the observed trends under status quo scenario. A. Covid-19 related ICU beds use B) Covid-19 related deaths C) Total number of individuals

## Highlights

In this paper, we used a stochastic, individual-based COVID-19 transmission model to evaluate the impact of the implemented social distancing interventions and to examine what would happen if those interventions had not been taken. We have shown that:

- If Greece had not implemented the social distancing interventions, the healthcare system would have been overwhelmed between 30 March and 4 of April.
- The early implementation of the social distancing measures would prevent 4360 deaths by 27 April
- Any interventions to boost ICU capacity, without the simultaneous implementation of SD interventions, is an inefficient healthcare policy, as the demand for those beds would be very high

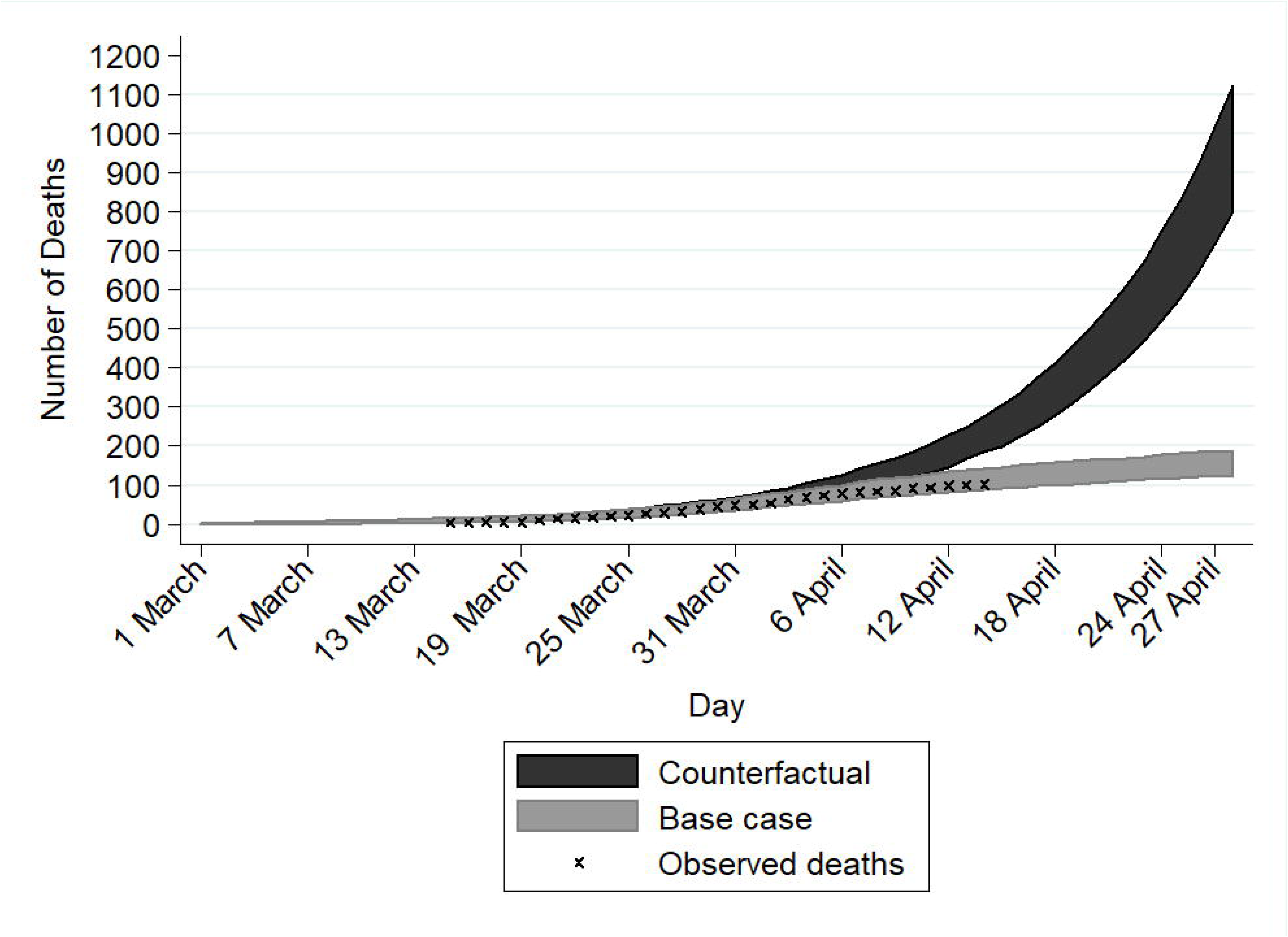

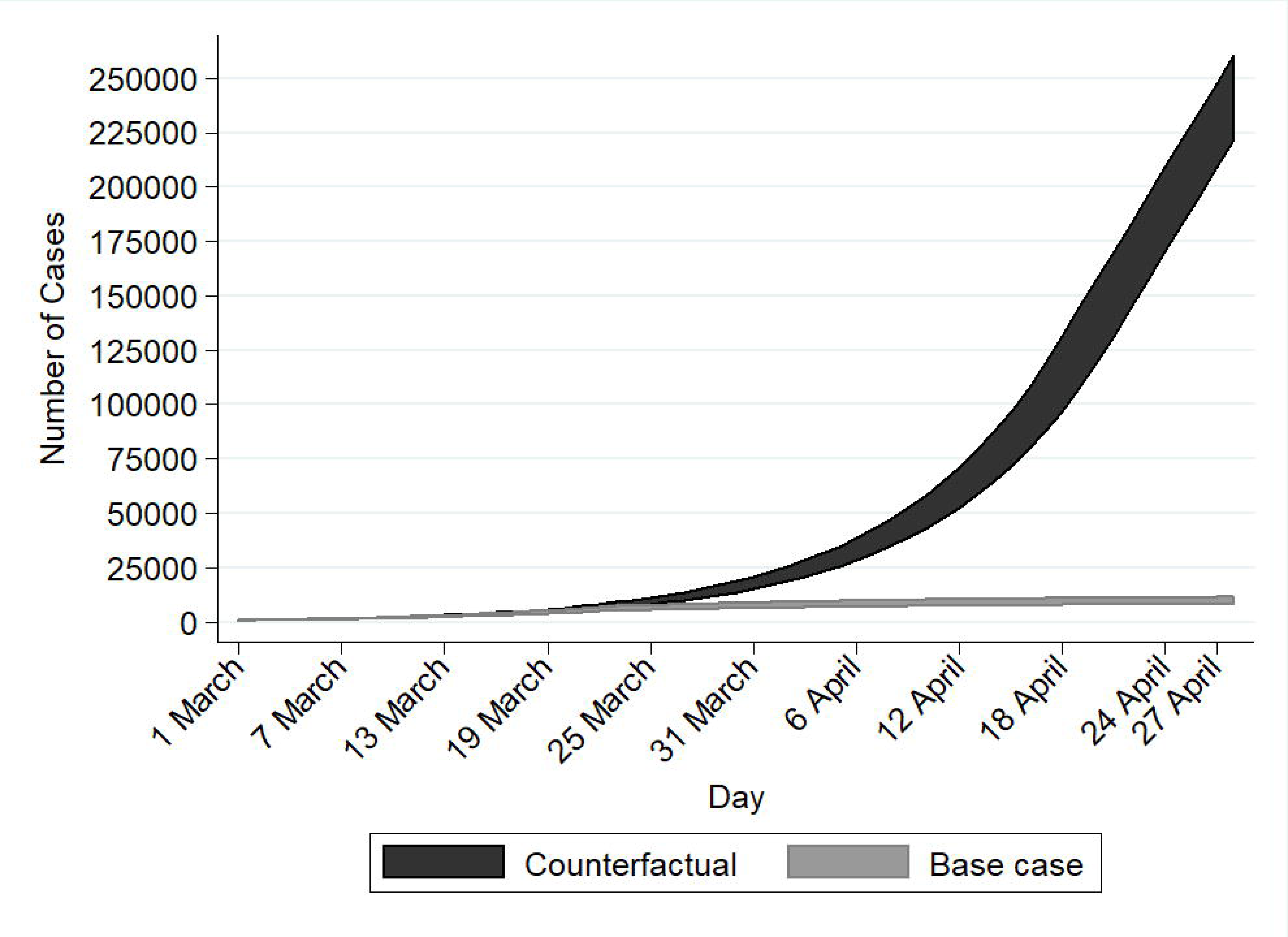

